# Integrating patient movement and pathogen genomics to support hospital infection prevention with PathoPath: a method development study

**DOI:** 10.64898/2026.06.03.26354630

**Authors:** Mohammad SI Sajib, Arif M Tanmoy, Naito Kanon, Anannya Barman Jui, Mohammad Shahidul Islam, Nigha Z Dola, Mohammad M Hossain, Reaz Mobarak, Mohammad Shahidullah, Mahbubul Hoque, ASM Nawshad Uddin Ahmed, Alison H Holmes, Samir K Saha, Senjuti Saha, Yu Wan, Yogesh Hooda

## Abstract

**Background:** Healthcare-associated infections pose a major burden to neonatal health worldwide and remain difficult to track in low-resource hospitals because patient movement data and pathogen genomic data are rarely integrated into actionable transmission models. Existing approaches are often restricted to specific settings, highly structured electronic health records (EHRs), or analyses focused on either patient movements or pathogen characteristics alone. To address this gap, we developed PathoPath, an open-source integrative modelling platform, and evaluated its utility in a high burden paediatric hospital in Dhaka, Bangladesh.

**Methods:** PathoPath is an open-source R package that combines electronic health records with whole genome sequencing data to generate contact networks from direct and indirect contacts using minimal structured inputs. We retrospectively applied PathoPath to 373 cases of *Klebsiella pneumoniae* species complex (KpSC) infection identified in 2021 at the largest paediatric referral hospital in Dhaka, Bangladesh. Ward level patient movement trajectories were used to reconstruct contact networks, and genomic data from isolates from children <60 days were integrated to identify probable dissemination of bacterial clones and antimicrobial resistance plasmids.

**Findings:** PathoPath identified 750 direct contacts among 317 patients, forming 25 connected components, with the largest including 93 patients. KpSC infections were identified across 21 of 37 wards, with the neonatal intensive care unit accounting for 77.9% of all cases. Integration of genomic and network data distinguished sustained clustering of ST147 from multiple probable inter-clonal dissemination events involving IncFII plasmids carrying blaNDM-5 and/or blaOXA-181 within ST16. Four dominant sequence types accounted for 65.6% of sequenced isolates, and carbapenemase genes were detected in 95.8%.

**Interpretation:** PathoPath reconstructs hospital-wide contact networks and integrates them with pathogen genomics to map probable dissemination of pathogens and antimicrobial resistance using minimal structured clinical data. It could support more targeted infection prevention and control in hospitals where granular digital records are not available.

**Funding:** Gates Foundation, National Institute of Health Research, Wellcome Trust, and the David Price Evans Endowment.

**Research in context:** *Evidence before this study:* We searched PubMed (pubmed.ncbi.nlm.nih.gov) and Semantic Scholar (www.semanticscholar.org) databases in May 2025 for literature about existing methods for modelling healthcare-associated transmission of infectious diseases. We identified at least 3 Bayesian, 1 SEIR (Susceptible, Exposed, Infectious, Recovered) and 33 other models/approaches such as StEP and outbreaker2 to support these efforts. 19/37 of these methods have demonstrated their capability to reconstruct transmission or detect outbreak with varying level of accuracy depending on the context and pathogen. However, most of these tools frequently rely on dense patient metadata (e.g., Radio Frequency Identification sensors), specific types of electronic healthcare records that might not be readily available across settings, or a specific spatial structure. Also, most of these tools either focus on patient records or genomic data so there was a need for a generalised, integrative framework that can bridge the gap between clinical records with minimal digital input and pathogen genomic information.

*Added value of this study:* We developed PathoPath, an open-access integrative pathogen agnostic method that uses minimal electronic healthcare data and genome information to construct transmission pathways of pathogen and antimicrobial resistance within a healthcare facility. We have tested this package using 373 cases of *Klebsiella pneumoniae* species complex (KpSC) infection at Bangladesh Shishu Hospital and Institute (BSHI), the largest tertiary paediatric healthcare facility in Dhaka which serves as a referral centre in Bangladesh for the sick paediatric population. Using PathoPath, we successfully identified sustained *K. pneumoniae* ST147 clonal transmission and a few events of potential dissemination of carbapenemase-encoding plasmids among *K. pneumoniae* ST16, providing us a better understanding on how resistance is moving through the hospital network.

*Implications of all the available evidence:* PathoPath provides a scalable approach to generate high resolution data to inform infection prevention and control in settings that still lack well-defined electronic healthcare records. This could provide a greater level of understanding of transmission than that offered by standard practice and enable the implementation of targeted, cost-effective interventions to prevent further outbreaks or spread of antimicrobial resistance within hospitals.

## Introduction

Healthcare-associated infections (HAIs) are a major cause of mortality and morbidity among neonates, especially in resource-constrained low-and-middle-income countries (LMICs).^1^ Limited staffing, overcrowding, and inadequate infection prevention and control (IPC) infrastructure allow sustained transmission of pathogens that are well adapted for such environment.^2^ The rapid emergence and dissemination of carbapenem resistance, particularly among *Klebsiella pneumoniae, Acinetobacter baumannii, Pseudomonas aeruginosa*, and *Enterobacteriaceae* species has further constrained effective therapeutic options, prompting the World Health Organisation (WHO) to classify these organisms as critical priority pathogens.^3^ Carbapenem resistant gram-negative bacterial clones and/or carbapenemase-encoding mobile genetic elements (MGEs) can spread rapidly within healthcare facilities through shared wards, equipment, healthcare worker or contaminated surfaces, making timely identification of transmission pathways essential for targeted IPC interventions.^4^

Several studies have utilised electronic healthcare records (EHRs), whole-genome sequencing (WGS), or both to reconstruct within/between healthcare facilities using Bayesian, agent-based or network modelling frameworks.^5-9^ These approaches have shown that combining patient movement data with pathogen genomics can substantially improve outbreak detection and transmission inference.^7,10^ However, many existing methods rely on highly structured digital health records, dense metadata, specific spatial structure, or advanced informatics systems^6,8^, which remain unavailable in many LMIC settings, where clinical documentation remains fragmented or paper-based.^11^ As a result, tools that operate with minimal structured clinical data remain scarce.

To address this gap, we developed PathoPath, an open-source pathogen agnostic R package that requires minimal EHR data and integrated them with WGS results to build a comprehensive contact network to help visualise potential transmission pathways of pathogen and/or MGEs to inform IPC strategies. As a proof-of-concept, we applied PathoPath to retrospective genomic and clinical data from *Klebsiella pneumoniae* species complex (KpSC) at a large paediatric referral hospital in Dhaka, Bangladesh.

## Methods

### Integrative modelling of contacts using PathoPath

We developed a generalised methodological framework to model spatiotemporal contacts between movement trajectories, expanding the target subjects from patients to include other individuals, and inanimate objects. The framework takes as input unique *subject* and *pathway* identifiers, along with date-stamped locations for each subject and pathway. For each patient in hospital settings, each pathway can be defined as a series of *locations* (bed, room, ward, *etc*, at the same granularity level) where the patient has stayed from admission to discharge. We have implemented this framework as an R package *pathopath* and released it on GitHub (github.com/wanyuac/pathopath). Here, we describe pathopath’s algorithm and analysis of contacts between the 373 inpatients admitted to the BSHI in 2021.

Assuming subject *s*_*1*_ entered and left location *l* on days *t*_*1s*_ and *t*_*1e*_ (*t*_*1s*_ ≤ *t*_*1e*_), respectively, and subject *s*_*2*_ entered and left the same location on days *t*_*2s*_ and *t*_*2e*_ (*t*_*2s*_ ≤ *t*_*2e*_), respectively, then days of s1 and s2’s stay at this location were determined by *L*_1_ *= t*_1*e*_ − *t*_1*s*_ + 1 for *s*_*1*_ and *L*_2_ *= t*_2*e*_ − *t*_2*s*_ + 1 for *s*_2_. The maximum length of days when subjects *s*_1_ or *s*_2_ or both were continuously present at location / was calculated as *L_max_* = *max* (*S*_1*e*_, *S*_2*e*_) − *min* (*S*_1*s*_, *S*_2*s*_)+ 1. Then we calculated the time interval *I*_12_ *= L_max_* − (*L*_1_ + *L*_2_).A direct contact between *s*_*1*_ and *s*_*2*_ at location / was identified when *I*_12_ < 0, and this contact had a length of *t*_*d*_ *=* −*I*_12_. Given a custom parameter *Δt* that represented a number of days, an indirect contact was identified when 0 < *I*_12_ < ∆*t*, and this contact had a length of *t*_*i*_ *=* ∆*t* − *I*_12_. The contact length was set to zero if neither direct nor indirect contact occurred between *s*_1_ and *s*_2_ at this location.

An undirected network was generated from all identified direct contacts. In this network, each node represented a patient, since each patient had a single movement pathway in our dataset, and each edge connected patients in at least one direct contact throughout their pathways, with the edge weight denoting the total length of direct contacts. Components were discovered in this contact network as subgraphs consisting of connected nodes that were not part of any larger connected subgraphs, and singletons were defined as individual nodes that did not connect to any other nodes. Bacterial species, genotypes, and SNP information were incorporated into the network as node attributes.

To track potential transmission, patients in the contact network were clustered using the same component-discovery algorithm but based on core-genome SNP distances between isolates recovered from these patients. Briefly, this method imports a matrix of pairwise core-genome SNP distances as an all-to-all undirected distance network, prunes edges with a configurable distance threshold, and identifies components of patients in the remaining distance network. Finally, such pathogen-based clusters were overlaid onto the contact network for comparison with the contact-based clusters. Readers are referred to **Supplementary appendix 1, page 11** for details.

### Study site and patient population

To assess the utility of PathoPath in a high-burden LMIC setting, this study was conducted at the BSHI which is the largest paediatric referral hospital in Dhaka, Bangladesh, with 37 unique wards having a total of 673-beds (**Supplementary appendix 1, Table S1, page 5**). We retrospectively retrieved EHRs and bacteriological stock cultures of all (n = 373) cases with culture-confirmed KpSC identified in 2021. While blood and/or cerebrospinal fluid (CSF) samples were routinely collected at BSHI for bacteriological culture, this study only included hospitalised children who met the WHO criteria for pneumonia, severe disease, or meningitis, assessed by trained physicians^14^. A detailed description of the bespoke EHR generation is described in **Supplementary appendix 1, page 9**.

### Ethical approval

This work was conducted under the ethics approval No. BICH-ERC-1/2/2018 from the Ethical Review Board of the Bangladesh Shishu Hospital and Institute (BSHI), formerly Bangladesh Institute of Child Health.^12,13^ Written informed consent was obtained from each patient’s legal guardian prior to data collection and analyses.

### Microbiological identification and whole-genome sequencing

The details of the microbiological identification are described in **Supplementary appendix 1, page 9**. Of the 373 culture-positive KpSC isolates identified in 2021, 314 isolates were from children aged <60 days. Of these, 311 (99.0%) isolates were successfully sequenced; three isolates from this age group could not be revived for sequencing. This age group was prioritised because it represents the population at highest risk for KpSC healthcare-associated infection in Bangladesh and accounted for the major burden of severe neonatal disease in this setting. Restricting sequencing to this high-risk subgroup also allowed more focused investigation of potential hospital transmission clusters. DNA extraction and 150-bp paired-end Illumina sequencing are described in **Supplementary appendix 1, pages 9–10**.

### Data analysis

Genomic typing including KL, OC, and MLST assignment was done on all sequenced KpSC isolates (**Supplementary appendix 1, page 10**).^15^ Steps involved in reference-based SNP calling and genomic analysis are provided in **Supplementary appendix 1, page 10**. For comparative plasmid analysis, plasmid sequences were reconstructed from draft genome assemblies (**Supplementary appendix 1, page 11**) and carbapenemase-encoding plasmid clusters were identified through a protocol adapted from a previous study (**Supplementary appendix 1, page 12**).^16^

All statistical analyses and visualisations were performed in R v4.3.1. with packages dplyr, tidyr, stringr, lubridate, and readxl/openxlsx. Phylogenetic trees were visualised using iTOL v7 (itol.embl.de).

## Results

### Study population for evaluating PathoPath

We evaluated PathoPath retrospectively using available data on KpSC from 2021, the major multidrug resistant cause of neonatal HAIs in Bangladesh. In 2021, a total of 21,301 patients were admitted at Bangladesh Shishu Hospital & Institute with an average of 1,775 admissions per month. Among 2,688 blood/CSF samples collected, 373 (13.9%) were culture-positive for KpSC and were recruited in this study. Patient demographics and outcomes are listed in Table 1. Most infections occurred early in life, with 47.2% (n = 177) presenting from primary clinics, birthing centres, and households across 27 districts in Bangladesh within the first week after birth, and 57.9% (n = 216) were hospitalised between 0 and 2 days before infection was identified (**Table 1**).

**Table 1:** Demographic and clinical characteristics of all 373 blood/CSF culture-positive KpSC cases at Bangladesh Shishu Hospital & Institute in 2021. Of the children older than 30 days, eight were aged <60 days (2 months). Of 314 infants aged <60 days, 311 (99.0%) were sequenced; three isolates in this group could not be revived. The 59 children aged ≥60 days were not sequenced.

| Patient population summary | Number of patients | Percentage (%) |
| --- | --- | --- |
| **Sex** |  |  |
| Female | 118 | 31.6 |
| Male | 255 | 68.4 |
| **Sample source** |  |  |
| Blood | 372 | 99.7 |
| Cerebrospinal fluid | 1 | 0.3 |
| **Outcome** |  |  |
| Left against medical advice | 40 | 10.7 |
| Died | 201 | 53.9 |
| Discharged | 132 | 35.4 |
| **Age (days) before infection identified** |  |  |
| 0–2 | 27 | 7.2 |
| 3–7 | 150 | 40.2 |
| 8–14 | 75 | 20.1 |
| 15–30 | 54 | 14.5 |
| 31–59 | 8 | 2.1 |
| ≥60 | 59 | 15.8 |
| **Hospital stay (days) before infection identified** |  |  |
| 0–2 | 216 | 57.9 |
| 3–7 | 115 | 30.8 |
| 8–14 | 26 | 7.0 |
| 15–30 | 14 | 3.8 |
| >30 | 2 | 0.5 |

### Hospital-wide contact network

Records of complete ward-level movement (from admission to discharge/death) were available for 317/373 (85%) patients and were used as input for Pathopath to reconstruct a hospital-wide network of direct contacts. This network comprised 750 direct contacts and 74 components (25 connected components and 49 singletons) (**Figure 1A**). The largest component consisted of 93 cases, accounting for 29.34% of the 317 KpSC-positive cases.

**Figure 1:**
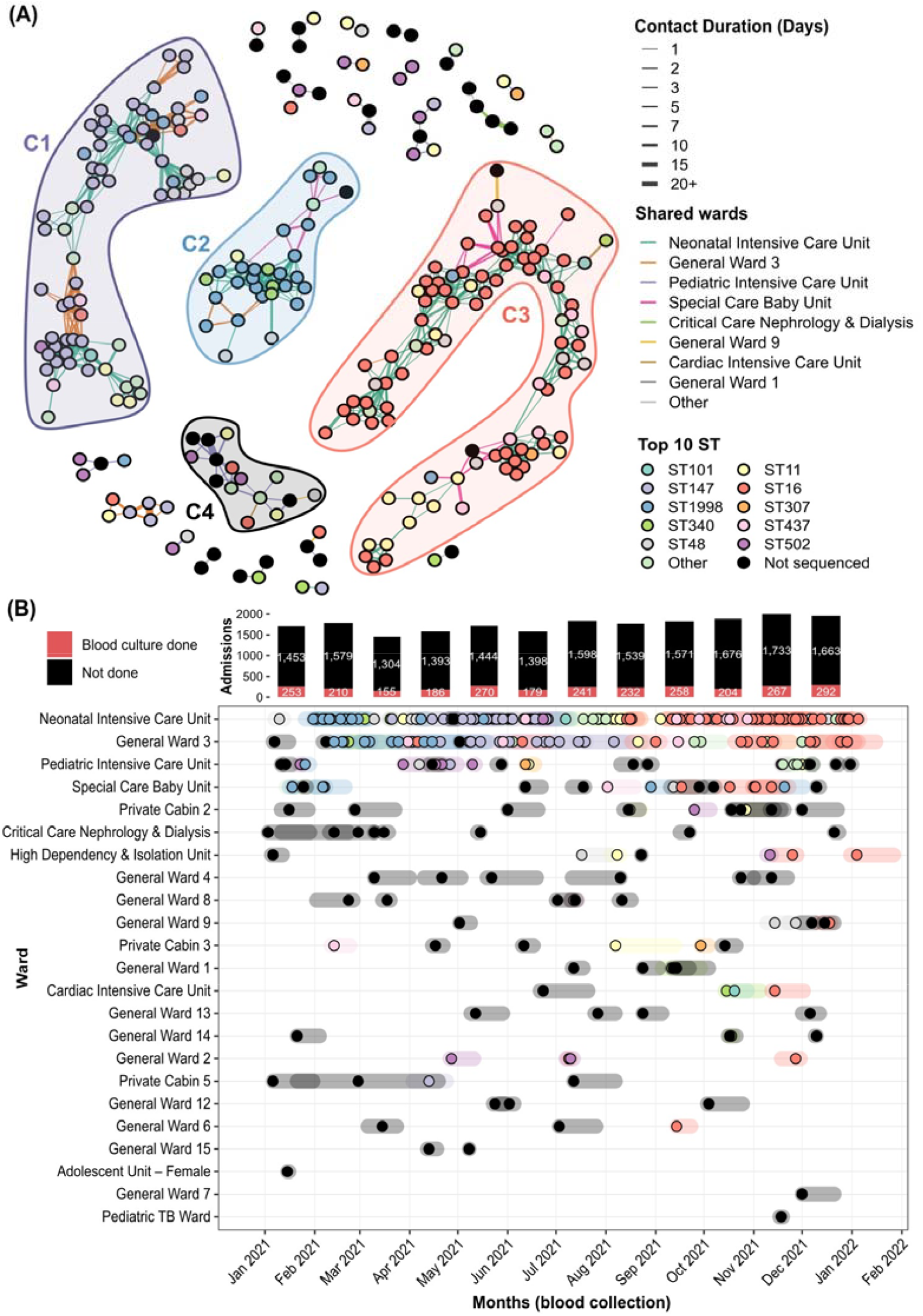
Complete ward-level contact network of KpSC culture-positive cases at BSHI in 2021. **(A)** Ward-level contact network of all 268 non-singleton KpSC isolates with nodes coloured by STs and edges indicating shared wards (where contacts occurred) and contact days between two indivuduals. The four major components C1-C4 are coloured by the top MLST. **(B)** Ward-level KpSC culture positive cases (n= 373) in 2021 with nodes coloured by STs. The coloured transparent bars across each nodes are highlighting admission to discharge dates for each individual. Total number of admissions and blood culture done at BSHI in 2021 are shown on the top bar chart.

KpSC infections were distributed across 21/37 (57%) hospital wards, but contact density was concentrated within neonatal critical care areas. The Neonatal Intensive Care Unit (NICU) had the highest burden of KpSC infections with approximately 77.9% of all contacts (584/750) involving 163 patients, followed by General Ward 3 (GW3: 10.3 % contacts from 47 patients), Special Care Baby Unit (SCBU: 25 cases and 5.3% of all contacts), and Paediatric Intensive Care Unit (PICU: 5.5% and 39 patients) (**Supplementary appendix 1, Table S4, page 7**).

### Genomic characterisation of KpSC isolates

Of the 373 archived KpSC isolates, 311 (83.4%) from children <60 days of age underwent WGS. These comprised 264 (85%) *K. pneumoniae* isolates of 24 MLST clones and 47 (15%) *K. quasipneumoniae* subsp. *similipneumoniae* isolates of 7 MLST clones (**Figure 1** and **Supplementary appendix 2, Sheet S1**). Four dominant STs accounted for 65.6% of sequenced isolates: ST16 (24.4%, n= 76), ST147 (18.7%, n = 58), ST1998 (12.2%, n = 38), and ST11 (10.3%, n = 32) (**Supplementary appendix 2, Sheet S2**).

Carbapenemase genes were detected in 298 (95.8%) isolates and were dominated by *bla*_NDM_ (n = 292) and *bla*_OXA_ (n = 99) genes followed by *bla*_KPC_ (n = 36). Notably, 42.1% (n = 130) isolates carried two carbapenemase genes. Specifically, 97.4% (74/76) of ST16 isolates harboured both *bla*_NDM-5_ and *bla*_OXA-181_, and only 2.6% (2/76) carried a single *bla*_OXA-181_. Approximately 98.3% (57/58) ST147 isolates carried *bla*_NDM-5_ and only one harboured the *bla*_NDM-1_ gene. For ST1998, the *bla*_KPC-2_ and *bla*_NDM-1_ combination was most frequent (86.8%; 33/38), followed by single *bla*_KPC-2_ (7.9%; 3/38), and *bla*_NDM-1_ (5.3%; 2/38). Carbapenemase genes in ST11 were mostly comprised of *bla*_NDM-7_ (19/32; 59·4%) and *bla*_NDM-5_ (13/32; 40·6%). Phenotypic resistance to Imipenem and meropenem were 98% for ST16, 100% for ST147 and 93.65% for ST1998 (**Table 2**). All the carbapenemase genes were harboured in plasmids, with IncFII (89.5% of ST16 and 93.1% of ST147), IncFIA(HI1)/IncFII (73.7% of ST1998), and IncFIB(K)/IncX3 (50% of ST11) being the most abundant vectors in our KpSC isolates (**Supplementary appendix 2, Sheet S3**).

**Table 2:**
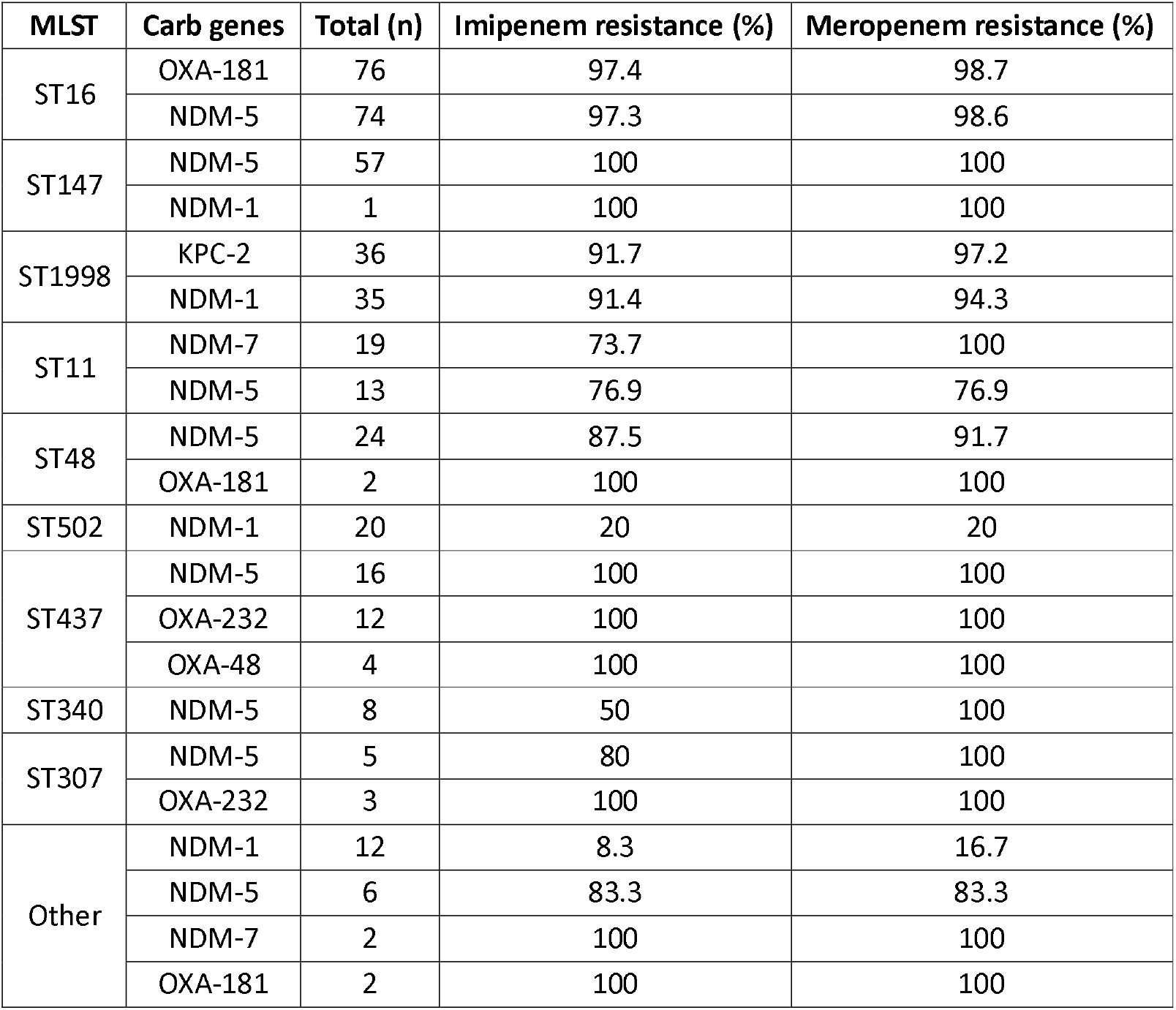
Carbapenem Resistance and associated genes by multi-locus sequence types (MLST) identified at BSHI.

#### Integration of genomic data into the contact network to infer transmission

Of the 317/373 cases with complete movement history, 263/317 were children <60 days age from whom sequencing data was available. The remaining 54 patients were either ≥60 days old or isolates could not be revived for sequencing. Once genotypical information was overlaid onto the network, >60% of cases on average within each of the three largest components (C1-C3) were dominated by a single ST (**Figure 1A**), K and O serotype (**Supplementary appendix 1, Figure S3, page 4; Table S5, page 8**). These major network clusters when combined with temporal trajectory of blood sample collection revealed concentrated seasonal waves of ST147 (C1), ST101 (C2) and ST16 (C3) mainly in NICU and GW3, the wards having the highest exchange of patients in between. These network patterns therefore prioritised ST147 and ST16, the largest two clusters (**Figures 1A, 1B**) for detailed analysis of clonal transmission and plasmid dissemination.

#### Identifying transmission clusters in ST147

All 58 ST147 KpSC cases at BSHI were detected between 18 March and 6 August 2021 (4.63 months), with an average of 2.88 cases per week (**Figure 1B**). Of these, 49/58 (84.5%) had complete ward-level movement records and therefore were used for the ST147-species network construction. PathoPath identified a total of 11 individual ST147 components linked by 110 edges (shared wards) and an average of 6.87 (± 5.82) pairwise contact days. Seven of the 11 components were singletons, with the largest network component having 29 patients connected (**Figure 2A**).

**Figure 2:**
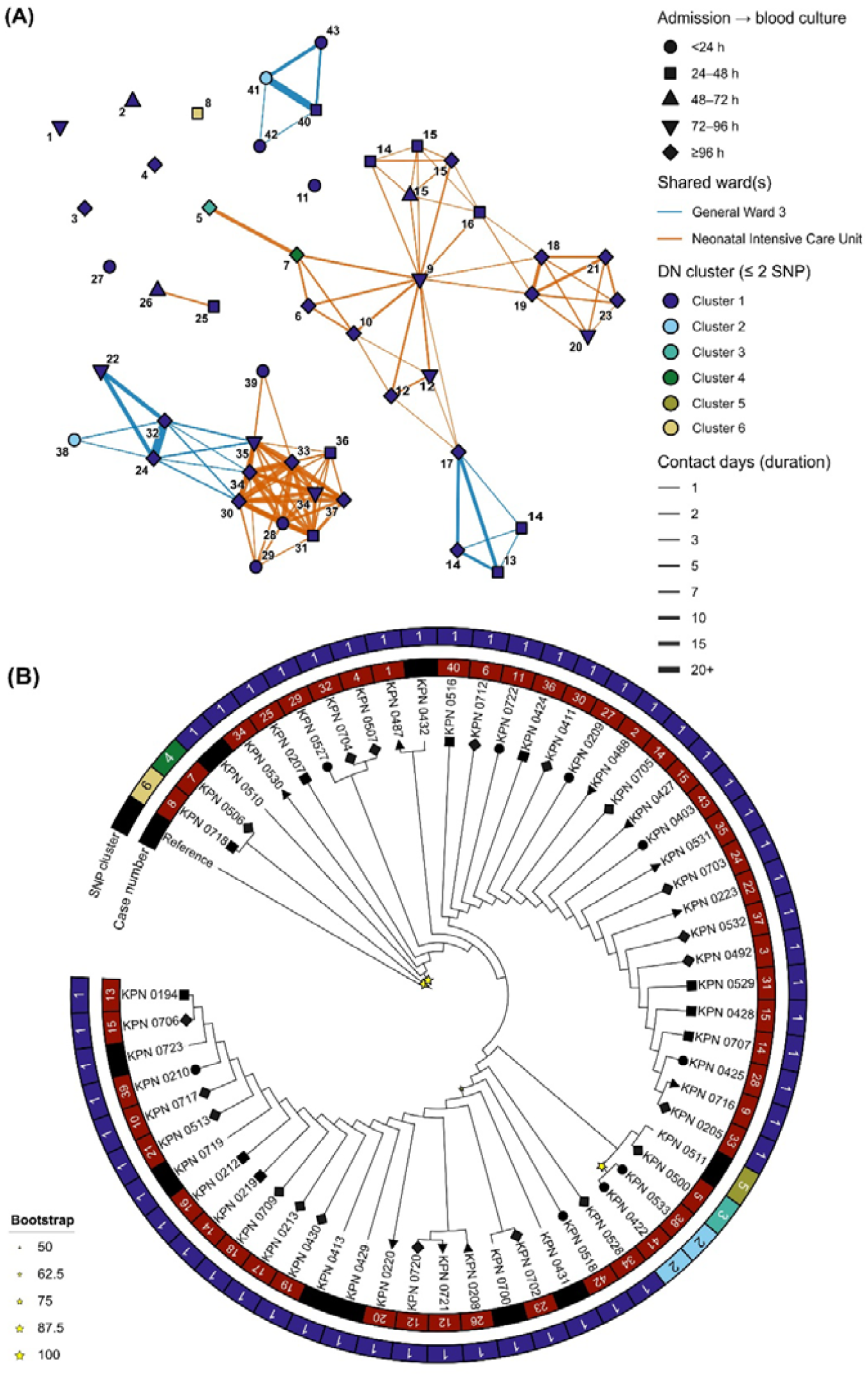
PathoPath contact network and maximum-likelihood (ML) phylogeny of KpSC ST147 culture-positive cases (n =58) at BSHI in 2021. **(A)** Ward-level contact network of all ST47 KpSC cases with nodes coloured by distance network clusters of single-nuceotide polymorphisms (SNPs) of isolates using a 2-SNP cutoff, and edges highlighting shared wards and contact days between two indivduals. Numbers on the nodes are highlighting chronological order of all ST147 cases detected at BSHI in 2021. Node shapes represent the time from admission to blood collection. **(B)** Unscaled single nuceotide polymorphism (SNP) based maximum-likelihood phylogenetic tree of ST147 with bootstap values (stars), and annotations showing SNP clusters and chronological order of cases (Case number). Tip shapes in the tree represent the time from admission to blood collection.

To identify DN clusters of potential transmission, genomic relatedness was assessed across a range of single nucleotide polymorphism (SNP) thresholds. Using a range of SNP distance cutoffs (0 to 100), between 3 and 21 distinct SNP clusters of ST147 were identified with an average of 5 partitions across all cutoffs. Overall number of partitions/clusters stabilised largely at >2 SNPs with no significant collapse for up to ≤100 SNP threshold tested (**Supplementary appendix 2, Sheet S4**). For further investigating potential transmission, we used ≤2 SNP as a threshold and found six distinct DN clusters. Notably, within Cluster 1, the largest of these six, 44/49 cases in the ST147 PathoPath network grouped together in just one compartment (DN Cluster 1), making this potential transmission lasting for about 141 days with an average number of 2.18 cases/week (**Figure 2A**).

Clinical timelines suggested mixed transmission timing within this dominant Cluster 1: 18 of 44 patients had blood cultures collected after at least 96 h of hospitalisation, whereas 16 had cultures drawn within 48 h of admission. Neonatal Intensive Care Unit (NICU), and General Ward 3 (GW3) were the only two wards where Cluster 1 infections were identified (NICU: 84 contacts involving 29 patients; GW3: 17 contacts involving 13 cases) (**Figure 2**). To account for environmental persistence and bridge any potential temporal gaps in transmission, a 28-day indirect contact window (Δt = 28) was applied. This created 255 additional contacts and included two singleton pairs, separated by 25 (Cluster 1) and 16 (Cluster 2) days. For both clusters, blood samples from the later patients were obtained on the day of admission.

#### Transmission of carbapenem resistance within ST16

Other than being the most abundant ST type (24.4.7%; 76/311) in our retrospective dataset, infections with ST16 at BSHI were continuously detected till the end of this study (from 6 April 2021 to 5 January 2022), with an average of 2.83 cases per week (**Figure 1B**). From this potentially ongoing transmission since 2021, 68/76 (89.5%) cases had complete movement history and thus were usable for constructing an ST16-specific network. Once built, a total of 12 components were identified with an overall 163 edges and approximately 3.52 (± 2.51) shared days on average between patients. There were nine singletons in the network plus three components with the largest SNP cluster of ST16 comprised of isolates from 47 cases (**Figure 3A**).

**Figure 3:**
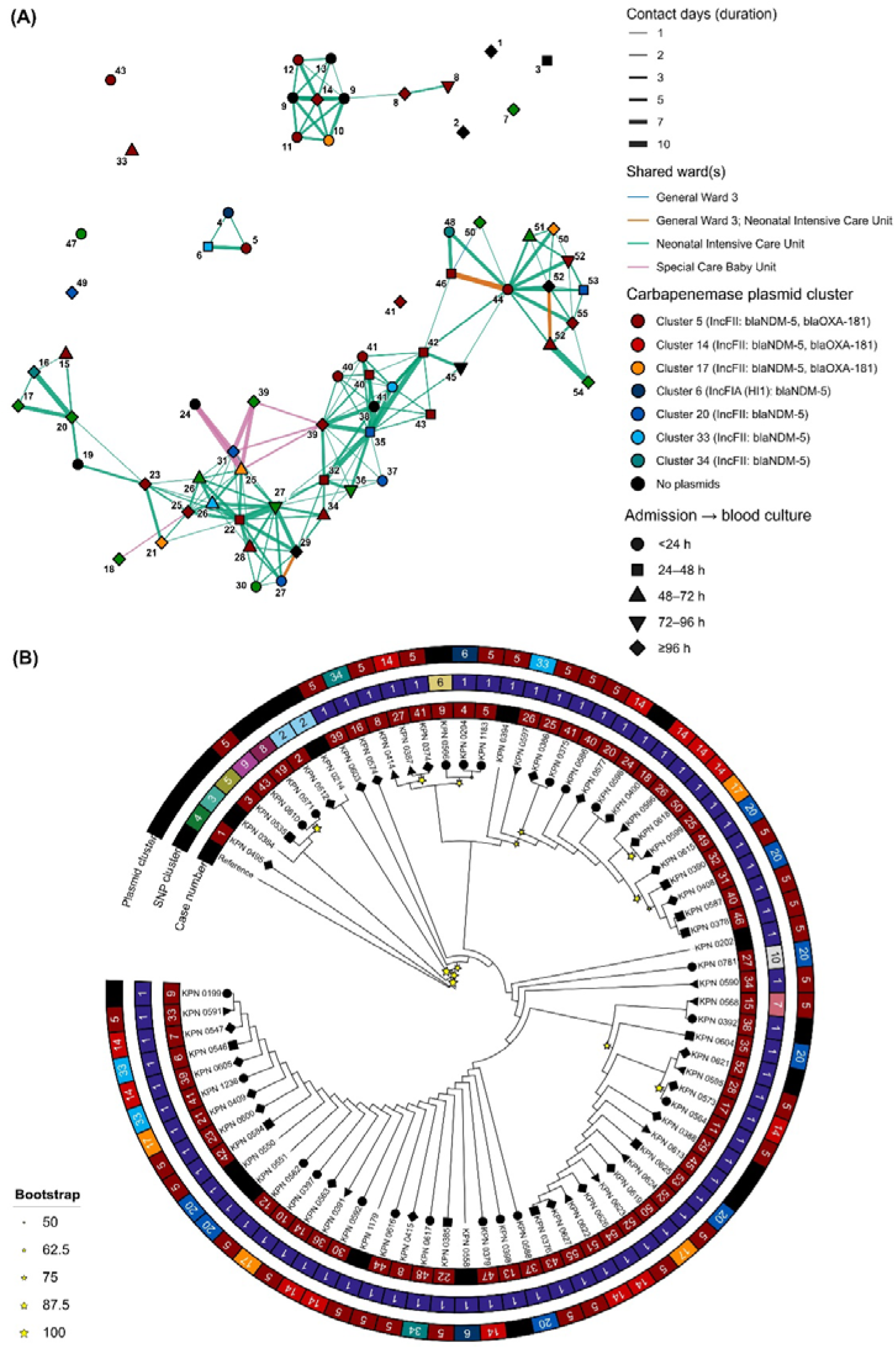
Integrative contact network and maximum-likelihood (ML) phylogeny of KpSC ST16 culture-positive isolates (n = 76) at BSHI in 2021. **(A)** Ward-level contact network of all ST16 KpSC isolates with nodes coloured by carbapenemase encoding plasmid clusters, AMR genes and replicon types, and edges highlighting shared wards and contact days between two indivuduals. Numbers on the nodes is highlighting chronological order of all ST16 cases detected at BSHI in 2021. **(B)** Unscaled single nuceotide polymorphism (SNP) based ML tree of ST147 with bootstap values (star), and annotations showing SNP/plasmids clusters and chronological order of cases. Shapes of the nodes in both network and ML tree are showing the time from admission to blood collection.

In total, 1 to 33 clusters of ST16 isolates were identified when varying the threshold of SNP distances from 0 to 100. The number and composition of clusters stabilised to less than 10 clusters when the distance threshold exceeded 2 SNPs (**Supplementary appendix 2, Sheet S4**). Therefore, the same ≤2 SNP threshold was used for clustering ST16 isolates as for the ST147, resulting in 10 clusters suggesting transmission of *K. pneumoniae* via direct contacts. Cluster 1, the largest continuous cluster had 69.1% (47/68) of all ST16 cases connected, spanning for 144 days with 2.92 cases/week until the end of this study.

Phenotypic resistance to carbapenems was nearly universal among ST16 as 97.4% (97.4% for *bla*_OXA-181_ and 97.3% for *bla*_NDM-5_) isolates were non-susceptible to imipenem and 98.7% (98.7% for *bla*_OXA-181_ and 98.6% for *bla*_NDM-5_) were resistant to meropenem (**Table 2**). Combining with the plasmid clusters, the contact network revealed potential inter-clonal transfer of carbapenemase-encoding plasmids (**Figure 3**). For example, an IncFII plasmid carrying *bla*_NDM-5_ (plasmid cluster 20) was observed to potentially move from a donor in the SNP Cluster 10 (patient 27) towards possibly a few SNP cluster 1 recipients (patients 31, 35, and 37) within the span of two weeks despite no recorded sharing of wards between these four patients, providing some insights gained to generate future research into environment or unrecognised transmission vectors. Similarly, an IncFII (cluster 5) plasmid carrying *bla*_NDM-5_ and *bla*_OXA-181_ genes potentially transferred from a group of SNP cluster 1 donors (patients 39, 40, 41, and 42) to a SNP cluster 9 recipient (patient 43) within the span of a week (**Figure 3**), suggesting horizontal dissemination of resistance determinants alongside clonal spread.

## Discussion

This study developed and tested PathoPath, an open-access, pathogen-agnostic tool that constructs hospital-wide contact network using only date-stamped patient movement trajectories and integrates genomic data to infer potential pathogen and/or AMR pathways. Using retrospective neonatal KpSC surveillance data from BSHI, we showed that PathoPath can identify dominant transmission clusters, precise pathways, and probable dissemination of carbapenemase-encoding plasmids within a high-burden tertiary hospital setting. Further ST-specific investigation revealed the value of this integrative framework as PathoPath delineated one transmission cluster of ST147, lasting for about 141 days in Neonatal Intensive Care Unit and General Ward 3. It also helped identifying transmission of IncFII carbapenemase-encoding plasmids (carrying *bla*_*NDM-5*_ and/or *bla*_*OXA-181*_ genes) between multiple SNP clusters of KpSC ST16 isolates. Together, these findings show how clonal expansion and plasmid-mediated resistance dissemination can coexist within the same healthcare network and remain difficult to detect without integrated digital surveillance.

PathoPath is designed to function with minimal structured clinical input. Because EHR in different healthcare facilities are often designed differently, including various commercial (e.g., Oracle Health EHR) or in-house data collection and management tools. Therefore, to standardise input structure of the patient movement trajectories a simplified table requiring subject identifier, pathway identifier, location, start time, and end time have been utilised in PathoPath. To minimise data-entry related errors in pathway reconstruction, the current version also includes a few logic checks, for example it flags patient overlaps (e.g., two patients occupying the same bed for multiple days) that are implausible. By demonstrating what can already be inferred from sparse routine data, PathoPath can in future help identify which additional metadata would be most informative to collect in future implementations. For example, interpretation of putative transmission events will always benefit from contextual information on time, place, and mechanism of exposure. In this sense, PathoPath can guide data-prioritisation efforts by showing where enhanced metadata collection, such as more granular timestamps, shared equipment records, healthcare worker movements, environmental sampling, or laboratory measures of pathogen persistence, would most improve transmission inference.

This is particularly relevant for genomic interpretation. While genomic clustering can be dynamically integrated using MLST, cgMLST, or LIN codes, SNP-based methods remain the most widely used approach for investigating hospital outbreaks because they offer the highest resolution for distinguishing closely related isolates.^15,17,18^ However, there is no universally appropriate SNP threshold for defining nosocomial transmission. A few important considerations to define nosocomial transmission of a single clone using SNPs is the geographical, temporal (typically 48-72 hours)^19^ and SNP distance (2-10 SNPs) thresholds, as arbitrary cutoffs may detrimentally affect the sensitivity and specificity of the inference.

Meaningful interpretation of contacts in depends on the interaction between genomic distance, temporal proximity, healthcare setting, and pathogen biology. For example, a recent multi-country study on *Klebsiella pneumoniae* systematically tested a varying range of temporal plus genetic distance cutoffs to identify the most sensitive and context-appropriate threshold (≤10 SNPs and ≤4 weeks).^17^ However, in our study, 26/76 (34.2%) ST16 isolates coming from 13 geographically distant districts were 0 SNPs apart from each other with a median admission to blood culture duration of 24 hours (mean = 3, min = 0 and max = 15 days). Although this finding should be interpreted cautiously and will require validation in future studies, it raises several possibilities. These include a shorter incubation period for ST16, particularly in neonates with underlying comorbidities, a slower rate of mutation accumulation than previously appreciated, which could suggest that these infections are part of a broader transmission network across Bangladesh, or transmission through contaminated hospital supplies, ambulances, or transport cots shared across healthcare facilities.^20^ For these reasons, we did not impose an absolute definition of HAI in this study and instead used ≤2 SNPs to make DN clusters as a pragmatic placeholder to demonstrate how PathoPath can support exploratory transmission mapping in both ST147 and ST16.

A similar principle applies to indirect contacts. PathoPath already allows users to model delayed or indirect interactions through a configurable Δt parameter, but in the present analysis this value was set to zero because the environmental persistence of KpSC under local hospital conditions remains uncertain. Experimental studies on environmental survival, ward-specific contamination, and clone-specific persistence could provide the empirical basis needed to calibrate indirect-contact windows more accurately.^21^ Likewise, prospective environmental sampling of high-contact surfaces, equipment, fluids, or shared clinical materials could clarify whether indirect transmission plays a substantial role in sustaining KpSC circulation in this setting.^22,23^ Such work would directly strengthen future implementations of PathoPath by enabling biologically informed parameterisation of contact networks.

While PathoPath is a scalable and versatile tool, it is important to mention a few limitations. First, the current network model is undirected and does not automatically infer transmission directionality. It is possible to utilise pathogen genome data and temporally infer directionality, our one-year sampling period did not support reliable evolutionary rate estimation. Second, although indirect contacts (Δt) can presently be modelled with PathoPath, uncertainty around environmental persistence prevented us from using non-zero Δt values in the retrospective BSHI analysis. Third, at BSHI, unique patient identifiers are not retained after discharge, which meant we could not link repeat admissions for the same individual across the study period. PathoPath can incorporate multiple pathways per patient, but this was not possible with the available dataset.

Despite these constraints, PathoPath offers a flexible digital framework where potential users can choose indirect interaction cutoffs based on their research context. Importantly, as open-source software, Pathopath enables local adaptation, providing users with method transparency and full control of code. While by integrating WGS information, PathoPath network can aid outbreak investigation, continuous genomic surveillance remains difficult to sustain. Therefore, future extensions of PathoPath may include the use of routinely available laboratory results in LMIC facilities (e.g., antibiotic zone diameters, minimum inhibitory concentration, and/or biochemical profiles), combined with machine learning tools to generate WGS-level inference and guide IPC practices in a more cost-effective and sustainable manner.

## Data Availability

All raw paired-end Illumina sequencing reads (fastq.gz) analysed and presented in this study are available on NCBI under the study accession "ERP173558" (Supplementary appendix 2, Sheet S1).
The PathoPath R package developed and used for modelling is openly accessible at: https://github.com/wanyuac/pathopath (Version 1.0.0; GPL-3.0 licence).

https://www.ebi.ac.uk/ena/browser/view/ERP173558

https://github.com/wanyuac/pathopath

## Data sharing

All raw paired-end Illumina sequencing reads (fastq.gz) analysed and presented in this study are available on NCBI under the study accession “ERP173558” (**Supplementary appendix 2, Sheet S1**).

The PathoPath R package developed and used for modelling is openly accessible at: https://github.com/wanyuac/pathopath (Version 1.0.0; GPL-3.0 licence).

## Declaration of interests

We declare no competing interests.

## Acknowledgments

The Authors acknowledge use of the High-Performance Computing platform provided by Liverpool Shared Research Facilities, Faculty of Health and Life Sciences, University of Liverpool.

## Funding

This work was funded by the NIHR Global Health Research Development Award (YH; Grant number: NIHR208273), the Gates Foundation (SS; Grant number: INV073135), and Wellcome Trust Centres for Antimicrobial Optimisation Network (CAMO-Net) Research Fellowship (MSIS; Grant number: 226691/Z/22/Z). YW is a Research Fellow funded by the David Price Evans Endowment to the University of Liverpool (Grant number: UGG0057). AHH is the David Price Evans Chair in Global Health and Infectious Diseases at the University of Liverpool (Grant number: UGG0057). AHH and YW were supported by the CAMO-NET UK grant (Reference: 226691/Z/22/Z).

## Supplementary materials

**Supplementary appendix 1**

**Supplementary appendix 2**

## Notes

### Competing Interest Statement

The authors have declared no competing interest.

### Author Declarations

This work was conducted under the ethics approval No. BICH-ERC-1/2/2018 from the Ethical Review Board of the Bangladesh Shishu Hospital and Institute (BSHI), formerly Bangladesh Institute of Child Health. Written informed consent was obtained from each patient's legal guardian prior to data collection and analyses.

## References

1. Allegranzi B, Bagheri Nejad S, Combescure C, Graafmans W, Attar H, Donaldson L, Pittet D. Burden of endemic health-care-associated infection in developing countries: systematic review and meta-analysis. Lancet 2011; 377(9761): 228–41.

2. Maki G, Zervos M. Health Care-Acquired Infections in Low- and Middle-Income Countries and the Role of Infection Prevention and Control. Infect Dis Clin North Am 2021; 35(3): 827–39.

3. Jesudason T. WHO publishes updated list of bacterial priority pathogens. The Lancet Microbe 2024; 5(9).

4. Dramowski A, Aucamp M, Beales E, et al. Healthcare-Associated Infection Prevention Interventions for Neonates in Resource-Limited Settings. Front Pediatr 2022; 10: 919403.

5. Pi L, Expert P, Clarke JM, Jauneikaite E, Costelloe CE. Electronic health record enabled track and trace in an urban hospital network: implications for infection prevention and control. 2021; 2021.

6. Pei S, Liljeros F, Shaman J. Identifying asymptomatic spreaders of antimicrobial-resistant pathogens in hospital settings. Proc Natl Acad Sci U S A 2021; 118(37).

7. Eyre DW, Laager M, Walker AS, Cooper BS, Wilson DJ, Program CDCMIDiH. Probabilistic transmission models incorporating sequencing data for healthcare-associated Clostridioides difficile outperform heuristic rules and identify strain-specific differences in transmission. PLoS Comput Biol 2021; 17(1): e1008417.

8. Didelot X, Kendall M, Xu Y, White PJ, McCarthy N. Genomic Epidemiology Analysis of Infectious Disease Outbreaks Using TransPhylo. Curr Protoc 2021; 1(2): e60.

9. Campbell F, Didelot X, Fitzjohn R, Ferguson N, Cori A, Jombart T. outbreaker2: a modular platform for outbreak reconstruction. BMC Bioinformatics 2018; 19(Suppl 11): 363.

10. Myall A, Price JR, Peach RL, et al. Prediction of hospital-onset COVID-19 infections using dynamic networks of patient contact: an international retrospective cohort study. Lancet Digit Health 2022; 4(8): e573–e83.

11. Woldemariam MT, Jimma W. Adoption of electronic health record systems to enhance the quality of healthcare in low-income countries: a systematic review. BMJ Health Care Inform 2023; 30(1).

12. Saha S, Saha SK. Invasive Bacterial Vaccine-Preventable Disease Surveillance: Successes and Lessons Learned in Bangladesh for a Sustainable Path Forward. J Infect Dis 2021; 224(12 Suppl 2): S293–S8.

13. Hooda Y, Tanmoy AM, Kanon N, et al. Escalating burden and mortality of carbapenem-resistant Klebsiella pneumoniae species complex infections in Bangladeshi infants. medRxiv 2025: 2025.06. 11.25329357.

14. Child WHODo, Health A. Handbook IMCI: integrated management of childhood illness: World Health Organization; 2005.

15. Lam MMC, Wick RR, Watts SC, Cerdeira LT, Wyres KL, Holt KE. A genomic surveillance framework and genotyping tool for Klebsiella pneumoniae and its related species complex. Nat Commun 2021; 12(1): 4188.

16. Lipworth S, Matlock W, Shaw L, et al. The plasmidome associated with Gramnegative bloodstream infections: A large-scale observational study using complete plasmid assemblies. Nat Commun 2024; 15(1): 1612.

17. Odih EE, Abdulahi J, Amulele A, et al. Contribution of nosocomial transmission to Klebsiella pneumoniae neonatal sepsis in Africa and South Asia: a meta-analysis of infection clusters inferred from pathogen genomics and temporal data. 2025.

18. Delgado-Blas JF, Rethoret-Pasty M, Brisse S. Life Identification Number (LIN) codes for the genomic taxonomy of Corynebacterium diphtheriae strains. Genome Med 2025; 18(1): 5.

19. Verani JR, Blau DM, Gurley ES, et al. Child deaths caused by Klebsiella pneumoniae in sub-Saharan Africa and south Asia: a secondary analysis of Child Health and Mortality Prevention Surveillance (CHAMPS) data. Lancet Microbe 2024; 5(2): e131–e41.

20. Lan HM, Lee YF, Tang YF, et al. Outbreak of Klebsiella aerogenes in a neurosurgical department linked to contaminated shampoo equipment from an outsourced barber department: A threat to infection control in outsourced healthcare services. J Microbiol Immunol Infect 2025.

21. Centeleghe I, Norville P, Hughes L, Maillard J-Y. Klebsiella pneumoniae survives on surfaces as a dry biofilm. American Journal of Infection Control 2023; 51(10): 1157–62.

22. Tofteland S, Naseer U, Lislevand JH, Sundsfjord A, Samuelsen Ø. A long-term low-frequency hospital outbreak of KPC-producing Klebsiella pneumoniae involving intergenus plasmid diffusion and a persisting environmental reservoir. PLoS One 2013; 8(3): e59015.

23. Hassan MZ, Sturm-Ramirez K, Rahman MZ, et al. Contamination of hospital surfaces with respiratory pathogens in Bangladesh. PLoS One 2019; 14(10): e0224065.

