## Supplementary appendix 1 for "Integrating patient movement and pathogen genomics to support hospital infection prevention with PathoPath: a method development study"

### Supplementary Figures

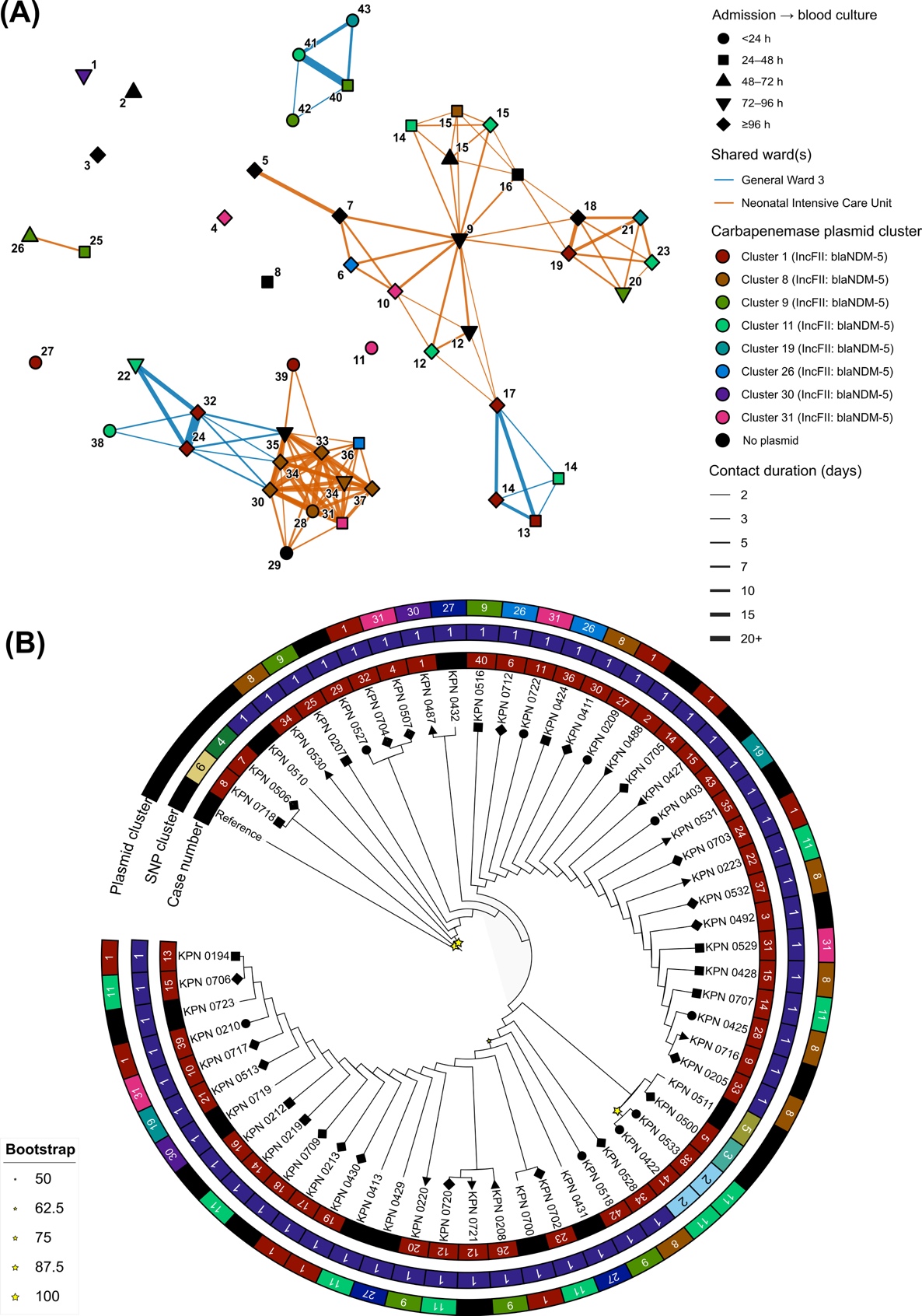

Figure S1: PathoPath contact network and maximum-likelihood (ML) phylogeny of KpSC ST147 **culture-positive isolates (n = 58) at BSHI in 2021. (A)** Ward-level contact network of all ST147 KpSC isolates with nodes coloured by carbapenemase encoding plasmid clusters and replicon types, and edges highlighting shared wards and contact days between two indivuduals. Number of node is highlighting chronological order of all ST147 cases detected at BSHI in 2021. **(B)** Unscaled single nuceotide polymorphism (SNP) based ML tree of ST147 with bootstap values (star), and annotations showing SNP/plasmids clusters and chronological order of cases. Shape of the nodes in both network and ML tree is showing the time from admission to blood culture.

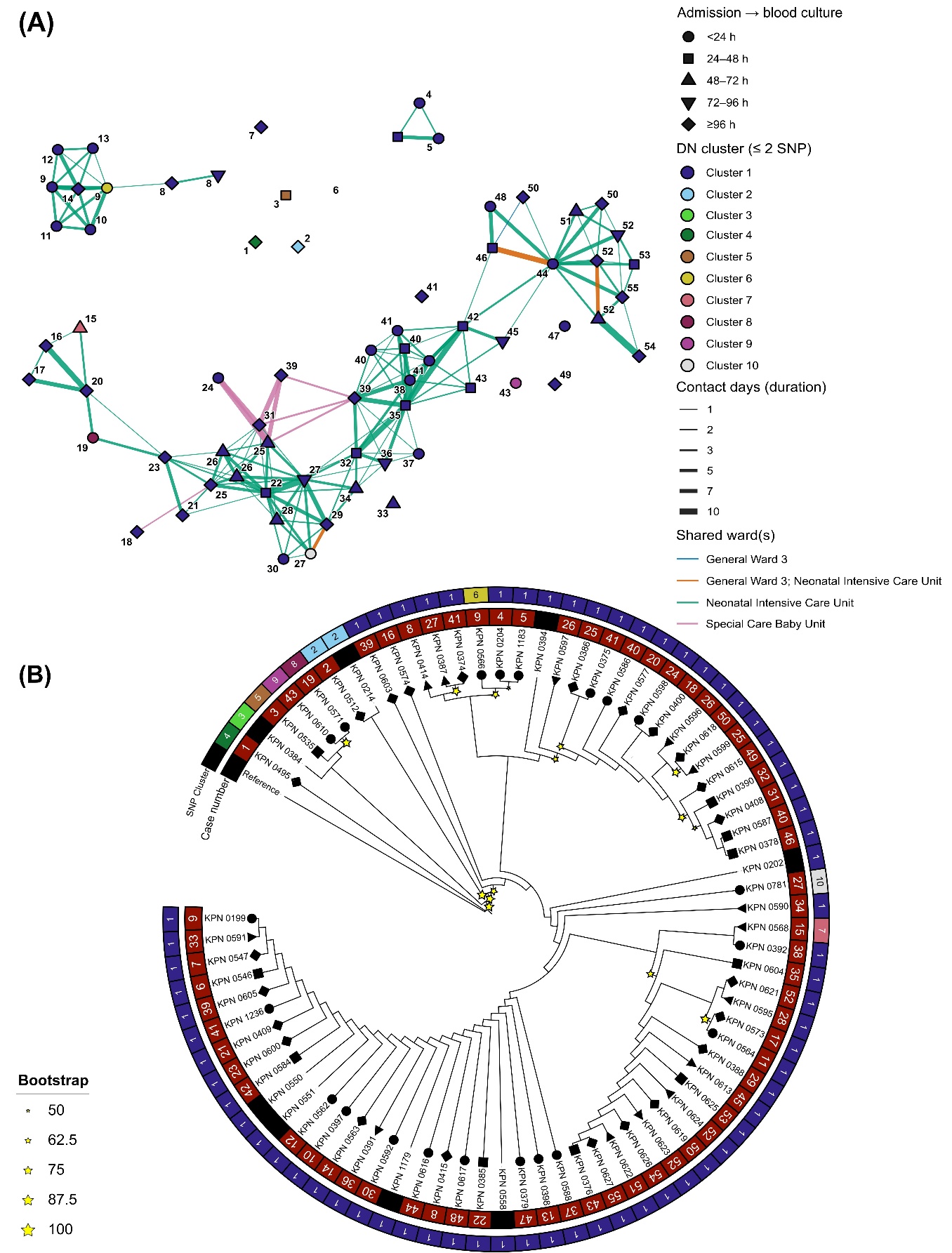

Figure S2: PathoPath contact network and maximum-likelihood (ML) phylogeny of KpSC ST16 **culture-positive isolates (n = 76) at BSHI in 2021. (A)** Ward-level contact network of all ST16 KpSC isolates with nodes coloured by single nuceotide polymorphism (SNP) based distance network (DN) clusters using 2 SNP cutoff, and edges highlighting shared wards and contact days between two indivuduals. Number of node is highlighting chronological order of all ST16 cases detected at BSHI in 2021. **(B)** Unscaled single nuceotide polymorphism (SNP) based ML tree of ST16 with bootstap values (star), and annotations showing SNP clusters and chronological order of cases (Case number). Shape of the nodes in both network and ML tree is showing the time from admission to blood culture.

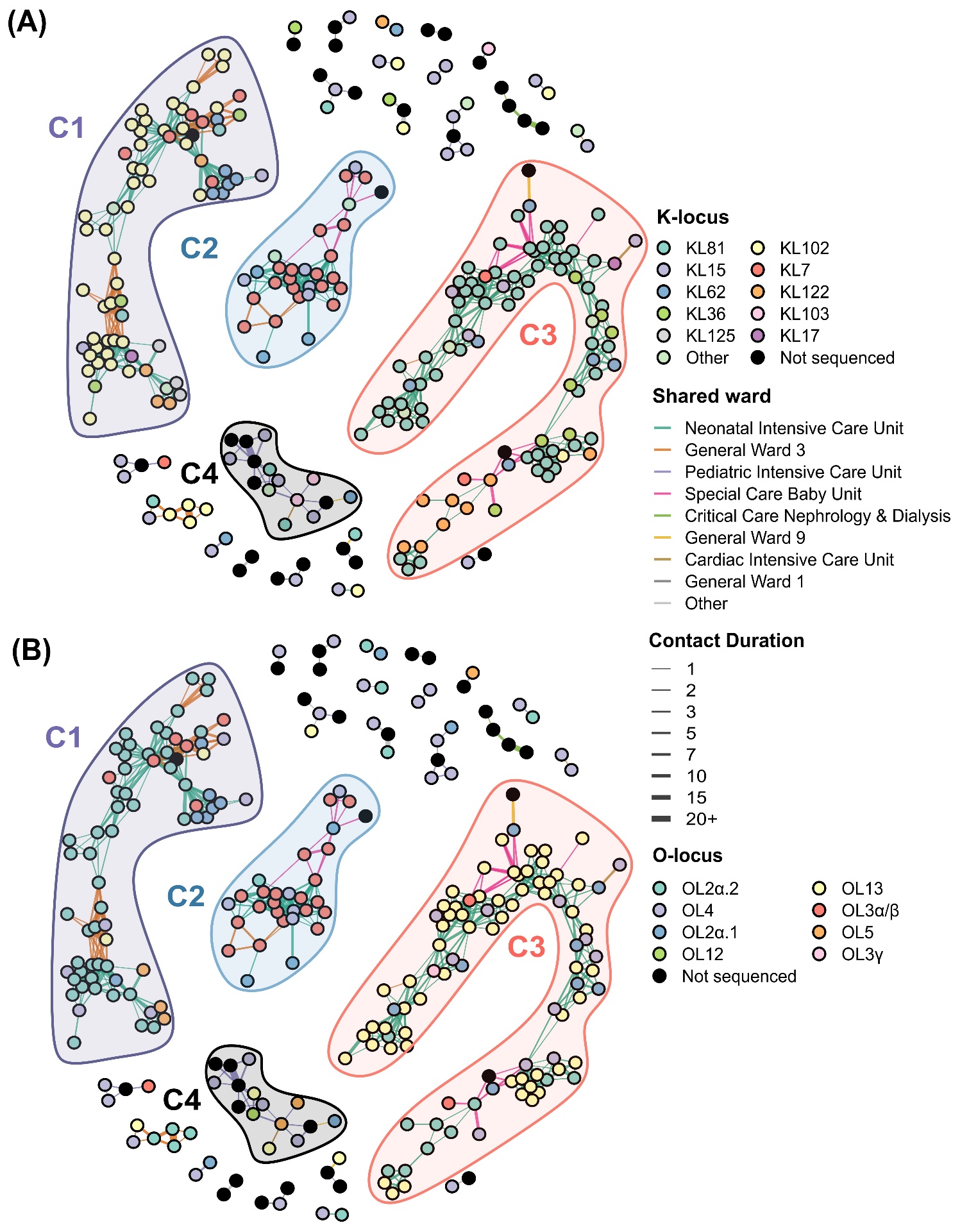

Figure S3: Complete PathoPath ward-level contact network of all non-singleton KpSC **culture-positive isolates (n = 268) at BSHI in 2021. (A)** Ward-level contact network of all KpSC isolates with nodes coloured by K-serotype. **(B)** Ward-level contact network of all KpSC isolates with nodes coloured by O-serotype. Coloured edges in the network is highlighting shared wards and contact length in days between two indivuduals.

### Supplementary Tables

#### Table S1: Structural layout of Bangladesh Shishu Hospital & Institute in 2021.

| **Block** | **Floor** | **Ward name** | **Layout type** | **No. of beds** |
| --- | --- | --- | --- | --- |
| Block A | Floor 1 | General Ward 1 | Open | 50 |
|  |  | General Ward 2 | Open | 42 |
|  |  | General Ward 7 | Open | 23 |
|  |  | High Dependency & Isolation Unit | Private room | 18 |
|  |  | Paediatric TB Ward | Open | 6 |
|  | Floor 2 | General Ward 3 | Open | 28 |
|  |  | Paediatric Diabetic Ward | Open | 6 |
|  |  | Special Care Baby Unit | Open | 18 |
|  |  | Neonatal Intensive Care Unit | Open | 16 |
|  |  | Paediatric Intensive Care Unit | Open | 16 |
|  |  | General Ward 4 | Open | 28 |
|  | Floor 3 | General Ward 5 | Open | 38 |
|  |  | General Ward 6 | Open | 53 |
|  |  | Paediatric Burn Unit | Open | 10 |
|  |  | Paediatric ENT Ward | Open | 2 |
|  |  | Private Cabin 2 | Private room | 24 |
|  |  | Adolescent Unit – Male | Open | 15 |
|  |  | Staff Health Unit | Private room | 6 |
|  | Floor 4 | Private Cabin 3 | Private room | 15 |
|  |  | Adolescent Unit – Female | Open | 15 |
| Block B | Floor 1 | General Ward 8 | Open | 26 |
|  |  | General Ward 9 | Open | 26 |
|  | Floor 2 | General Ward 10 | Open | 25 |
|  |  | General Ward 11 | Open | 12 |
|  | Floor 3 | Critical Care Nephrology & Dialysis | Open | 8 |
|  |  | General Ward 12 | Open | 13 |
|  |  | General Ward 13 | Open | 12 |
|  |  | Paediatric Renal & Dialysis | Cubical | 6 |
|  | Floor 4 | General Ward 14 | Open | 24 |
|  |  | General Ward 15 | Open | 17 |
|  | Floor 5 | Cardiac High Dependency Unit | Open | 5 |
|  |  | Cardiac Intensive Care Unit | Open | 7 |
|  |  | Cardiac Pre-Cath Unit | Open | 8 |
|  |  | Cardiac Post-Cath | Open | 6 |
|  | Floor 6 | Private Cabin 5 | Private room | 19 |
| Block C | Floor 3 | Cardiac Pre-op Holding | Open | 10 |
| Block D | Floor 1 | Paediatric COVID-19 Ward | Private room | 20 |

Table S2: Summary statistics generated with BUSCO for 311 draft genome assemblies. Abbreviation: SD, standard deviation.

| **BUSCO Metric** | **Mean** | **SD** | **Median** | **Minimum** | **Maximum** |
| --- | --- | --- | --- | --- | --- |
| **Complete** | 98.59 | 1.02 | 98.7 | 87.8 | 99.1 |
| **Duplicated** | 0.53 | 0.07 | 0.5 | 0.2 | 0.9 |
| **Fragmented** | 0.29 | 0.62 | 0.2 | 0 | 7.3 |
| **Missing** | 1.12 | 0.47 | 1.1 | 0.8 | 5.4 |
| **Single copy** | 98.07 | 1.01 | 98.2 | 87.3 | 98.6 |
| **Lineage** | 10 | 0 | 10 | 10 | 10 |
| **Total benchmark genes** | 440 | 0 | 440 | 440 | 440 |

#### Table S3: Summary statistics generated for 311 draft genome assemblies. Abbreviation: SD, standard deviation.

| **CheckM2 Metric** | **Mean** | **SD** | **Median** | **Minimum** | **Maximum** |
| --- | --- | --- | --- | --- | --- |
| **Average gene length (bp)** | 308 | 5 | 307 | 269 | 318 |
| **Coding density** | 0.88 | 0 | 0.88 | 0.87 | 0.89 |
| **Completeness** | 99.98 | 0.28 | 100 | 95.75 | 100 |
| **Contamination (%)** | 0.23 | 0.24 | 0.16 | 0 | 1.57 |
| **Contig (N50)** | 232,398.03 | 94,493.29 | 257,282.00 | 9,670.00 | 507,162.00 |
| **GC content (%)** | 0.57 | 0 | 0.57 | 0.56 | 0.58 |
| **Genome size (bp)** | 5,614,979 | 115,033 | 5,594,905 | 5,208,129 | 5,862,066 |
| **Max contig length (bp)** | 533,792 | 206,075 | 544,003 | 74,744 | 1,179,968 |
| **Total coding sequences** | 5,348.44 | 150.31 | 5,350.00 | 4,869.00 | 5,840.00 |
| **Total contigs** | 152.37 | 89.66 | 133 | 37 | 969 |

Table S4: Summary of all non-singleton ward-level patient contacts at BSHI by KpSC sequence types determined through multi-locus sequence typing (MLST).

| **MLST** | **Shared wards** | **No of contacts** | **No of patients** | **Total contact duration (days)** | **Mean days** |
| --- | --- | --- | --- | --- | --- |
| ST11 | Neonatal Intensive Care Unit | 11 | 10 | 32 | 2.91 |
| ST16 | Neonatal Intensive Care Unit | 153 | 56 | 670 | 4.38 |
|  | Special Care Baby Unit | 9 | 7 | 65 | 7.22 |
|  | General Ward 3 | 4 | 7 | 25 | 6.25 |
| ST48 | Neonatal Intensive Care Unit | 11 | 7 | 67 | 6.09 |
| ST147 | Neonatal Intensive Care Unit | 88 | 31 | 672 | 7.64 |
|  | General Ward 3 | 22 | 15 | 194 | 8.82 |
| ST334 | Paediatric Intensive Care Unit | 1 | 2 | 3 | 3 |
| ST437 | Neonatal Intensive Care Unit | 4 | 6 | 11 | 2.75 |
| ST340 | Neonatal Intensive Care Unit | 3 | 3 | 72 | 24 |
|  | General Ward 3 | 1 | 2 | 35 | 35 |
| ST378 | Neonatal Intensive Care Unit | 1 | 2 | 8 | 8 |
| ST502 | Paediatric Intensive Care Unit | 2 | 4 | 5 | 2.5 |
| ST1998 | Neonatal Intensive Care Unit | 38 | 17 | 223 | 5.87 |
|  | General Ward 3 | 8 | 9 | 65 | 8.12 |
|  | Special Care Baby Unit | 5 | 6 | 23 | 4.6 |
| Not sequenced | Paediatric Intensive Care Unit | 6 | 8 | 48 | 8 |
|  | Nephrology & Dialysis | 3 | 4 | 31 | 10.33 |
|  | Private Cabin 2 | 1 | 2 | 26 | 26 |
|  | Private Cabin 3 | 1 | 2 | 9 | 9 |
|  | General Ward 12 | 1 | 2 | 5 | 5 |

Table S5: Summary of top four ward-level patient contact components at BSHI.

| **Components (top 4)** | C1 | C2 | C3 | C4 |
| --- | --- | --- | --- | --- |
| **Component size** | 93 | 71 | 30 | 15 |
| **Dominant ST** | ST16 | ST147 | ST1998 | Not sequenced |
| **Dominant ST (N)** | 59 | 39 | 20 | 5 |
| **Dominant ST (%)** | 63.4 | 54.9 | 66.7 | 33.3 |
| **Dominant K** | KL81 | KL102 | KL7 | Not sequenced |
| **Dominant K (N)** | 59 | 41 | 19 | 5 |
| **Dominant K (%)** | 63.4 | 57.7 | 63.3 | 33.3 |
| **Dominant O** | OL13 | OL2α.2 | OL3α/β | Not sequenced |
| **Dominant O (N)** | 59 | 46 | 20 | 5 |
| **Dominant O (%)** | 63.4 | 64.8 | 66.7 | 33.3 |

### Methods

#### Generation of electronic health records

Demographic (age and sex) and clinical data, including symptoms, admission date, ward location and date of sample collection, were prospectively recorded at enrolment from all eligible patients using Android tablet–based surveillance platform developed by the Child Health Research Foundation (CHRF). During hospital stay, trained research physicians visited each enrolled patient daily to document medications administered, treatment modifications, bed or ward transfers, interim clinical changes, final diagnosis, and hospital outcomes, including discharge, death, or leaving against medical advice. Data were entered in real time using structured electronic forms with built-in logic checks to minimise entry errors and maintain consistency across variables. Records were subsequently curated centrally through the CHRF surveillance data system, where patient identifiers, admission trajectories, microbiological records, and movement histories were harmonised to generate analysable electronic health records (EHRs) for network modelling. For PathoPath analysis, only unique patient identifiers, location history, and date-stamped admission and transfer records were required.

#### Microbiological identification and whole-genome sequencing

For microbiological diagnosis, one blood/CSF specimen was collected from each case and cultured at 37°C using automated BACTEC systems (Becton Dickinson, USA) for up to 120 hours. Samples positive for microbial growth detected by the system were plated on blood, chocolate, and MacConkey agar media to isolate pure colonies. Next, bacterial genus and species were identified using standard biochemical tests (e.g., oxidase, indole, urease, citrate). Isolates with a oxidase-negative, indole-negative, urease-positive, and citrate-positive biochemical profile were identified as *Klebsiella*, as described in our previous study^1^. Pure colonies, one per patient, were stored at −80 °C in Skim milk–Tryptone–Glucose–Glycerol (STGG) media.

314/373 culture positive KpSC isolates, only from children <60 days of age, were chosen retrospectively for whole-genome sequencing (WGS), although 311/314 could be sequenced as three isolates could not be revived. This age group represents the highest-risk population for KpSC HAI in Bangladesh. Frozen stocks of KpSC isolates were subcultured onto MacConkey agar and incubated at 37°C overnight. Genomic DNA was extracted using QIAamp DNA Mini Kit (Qiagen, Germany; Cat. 56304) and DNA libraries were prepared with using the NEBNext Ultra II FS DNA Library Kit (New England Biolabs, USA; Cat. E7805). The WGS was performed with a 150-bp paired-end layout on an Illumina NextSeq 2000 platform (Illumina Inc., USA) at the Child Health Research Foundation (CHRF) in Dhaka.

#### KL, OC, and MLST assignment

Using fastp v1.0, raw Illumina reads were trimmed to remove sequencing adapters and poor-quality tails (average quality below Phred Q30 in a 4-bp sliding window) and were filtered to remove trimmed reads with less than 60% bases of a minimum quality Q30^2^. Draft genome assemblies were generated *de novo* from the processed reads using Unicycler (v0.5.1)^3^. The quality of each assembly was assessed using BUSCO (busco.ezlab.org/) and CheckM2 (github.com/chklovski/CheckM2) for a total contig length <7 Mbp, contig number <1000, and contamination level <5%. The capsular locus (KL), O-antigen cluster (OC), and multi-locus sequence type (MLST; Pasteur scheme)^4^ of each isolate were determined from the assembled contigs using Kleborate v3^5^.

#### *De novo* genome assembly, SNP calling and phylogenomic analysis

For phylogenomic analysis, ST16 and ST147 draft assemblies were separated in two groups and mapped against closely related ST-specific complete chromosome sequences (NZ_CP186626.1: ST16 and NZ_CP171583.1: ST147) to produce core-genome alignments using Snippy v4.6.0^6^. Repetitive regions (≥90% nucleotide identity) in each reference genome were determined using NUCmer (MUMmer v4)^7^ and scripts filterCoords.py and mergeGenomicRegions.py from the cgSNPs package (github.com/wanyuac/cgSNPs). Prophage regions were identified using PHASTEST v1.1^8^, and the resulting coordinates were masked from the core-genome alignments using BEDTools v2.31.1^9^. Recombination was detected and filtered out using Gubbins v2.4.1^10^ with 20 iterations and SNP-only sites after dropping invariants were extracted using snp-sites v 2.5.1^11^. Maximum-likelihood phylogenies were then generated with IQ-TREE2 v2.4.0^12^ using ModelFinder and 1,000 ultrafast bootstraps and pairwise SNP distances were calculated with snp-dists v.0.8.2^13^. Finally, to infer hierarchical Bayesian clusters, FastBAPS (R package v1.0.8)^14^ was applied to the repeat/phage/recombination-filtered core SNP alignments of the two dominant MLST types (ST16 and ST147) in our dataset.

#### Clustering of patients by pathogen similarity

Pathopath has implemented two approaches to patient clustering on the basis of pathogen similarity. The first approach, as demonstrated in this study, discovers components in a distance network of pathogens that were recovered from patients in the contact network. This approach accounts for cumulative micro-evolutionary drift and groups closely related isolates into potential nosocomial transmission chains. It has been implemented in PathoPath as function dn_clustering. This function imports a matrix of pairwise core-genome single nucleotide polymorphism (SNP) distances, which is equivalent to an all-to-all undirected graph where nodes represent individual sequenced isolates/clones and edge weights are SNP distances. Then entries greater than a configurable threshold were masked in the matrix, corresponding to pruning edges to retain those with weights smaller or equal to the threshold$.$ Finally, using the igraph^15^ package in R, discrete connected components within the remaining network are identified and assigned a unified integer cluster membership ID and a corresponding display label (e.g., "Cluster 1", "Cluster 2"), which are exported as a structured data frame (tibble) to be overlaid onto the contact network.

The second approach, which has been implemented in Pathopath as function h_clustering, performs complete-linkage hierarchical clustering on the distance matrix and cuts the resulting dendrogram into clusters where no pairwise distance exceeds the given threshold. By definition, result of this approach converges with the component-discovery approach (dn_clustering) when the distance threshold equals zero, reporting clusters of identical isolates.

#### Comparative plasmid analysis

Plasmids were reconstructed from draft genome assemblies using the mob_recon tool of MOB-suite v3.1.5, with contigs filtered in the context of Enterobacteriaceae chromosomes (zenodo.org/records/3785351, v2019-11-NCBI) to improve accuracy. Replicon types of reconstructed plasmids were screened against the PlasmidFinder database (updated on 20 August 2025) using ABRicate v1.0.1 (github.com/tseemann/abricate) for ≥85% nucleotide identities and ≥90% coverages. Plasmid-borne AMR genes were detected using AMRFinderPlus v4.0.23 and its database v2025-07-16.1.

We followed the method developed by Matlock, Lipworth, Chau, *et al* to identify clusters of carbapenemase-encoding plasmids^16^. Specifically, all-to-all genetic distances were calculated from reconstructed plasmids using MASH v2.3 (sketch size: one million; k-mer size: 21 bp) and were converted to similarities by subtracting from 1. The pairwise length ratio of these plasmids were calculated with the equation $min(L_{i},L_{j})/max(L_{i},L_{j})$, where *L_i_* and *L_j_* represent the lengths of two plasmids. A network was generated for plasmids showing genetic similarities >0.9999 and length ratios >0.99 and was visualised in Cytoscape v3.10.3 (cytoscape.org). Identification of plasmid clusters was performed based on the genetic similarities using the Louvain method as implemented in function cluster_louvain of the igraph package in R.

#### Data visualisation and statistical analysis

All statistical analysis and data visualisation were performed in R v4.3.1. Metadata or genome information were loaded, cleaned, reshaped, or summarised using R packages dplyr, tidyr, stringr, lubridate, and readxl/openxlsx. Phylogenetic and data visualisation was done using iTOL v7^17^ and R packages ggplot2, ggrepel, and tidyverse.

### References

1.	Hooda Y, Tanmoy AM, Kanon N, et al. Escalating carbapenem-resistant Klebsiella burden and mortality in Bangladeshi infants. *medRxiv* 2025: 2025.06. 11.25329357.

2.	Chen S. fastp 1.0: An ultra-fast all-round tool for FASTQ data quality control and preprocessing. *Imeta* 2025; **4**(5): e70078.

3.	Wick RR, Judd LM, Gorrie CL, Holt KE. Unicycler: Resolving bacterial genome assemblies from short and long sequencing reads. *PLoS Comput Biol* 2017; **13**(6): e1005595.

4.	Diancourt L, Passet V, Verhoef J, Grimont PA, Brisse S. Multilocus sequence typing of Klebsiella pneumoniae nosocomial isolates. *J Clin Microbiol* 2005; **43**(8): 4178-82.

5.	Lam MMC, Wick RR, Watts SC, Cerdeira LT, Wyres KL, Holt KE. A genomic surveillance framework and genotyping tool for Klebsiella pneumoniae and its related species complex. *Nat Commun* 2021; **12**(1): 4188.

6.	Seemann T. Snippy: Rapid haploid variant calling and core genome alignment.; 2020.

7.	Marcais G, Delcher AL, Phillippy AM, Coston R, Salzberg SL, Zimin A. MUMmer4: A fast and versatile genome alignment system. *PLoS Comput Biol* 2018; **14**(1): e1005944.

8.	Wishart DS, Han S, Saha S, et al. PHASTEST: faster than PHASTER, better than PHAST. *Nucleic Acids Res* 2023; **51**(W1): W443-W50.

9.	Quinlan AR. BEDTools: The Swiss-Army Tool for Genome Feature Analysis. *Curr Protoc Bioinformatics* 2014; **47**: 11 2 1-34.

10.	Croucher NJ, Page AJ, Connor TR, et al. Rapid phylogenetic analysis of large samples of recombinant bacterial whole genome sequences using Gubbins. *Nucleic Acids Res* 2015; **43**(3): e15.

11.	Page AJ, Taylor B, Delaney AJ, Soares J, Seemann T, Keane JA, Harris SR. SNP-sites: rapid efficient extraction of SNPs from multi-FASTA alignments. *Microb Genom* 2016; **2**(4): e000056.

12.	Minh BQ, Schmidt HA, Chernomor O, Schrempf D, Woodhams MD, von Haeseler A, Lanfear R. IQ-TREE 2: New Models and Efficient Methods for Phylogenetic Inference in the Genomic Era. *Mol Biol Evol* 2020; **37**(5): 1530-4.

13.	Seemann T. SNP-dists: Pairwise SNP distance matrix from a FASTA sequence alignment. 2021.

14.	Tonkin-Hill G, Lees JA, Bentley SD, Frost SDW, Corander J. Fast hierarchical Bayesian analysis of population structure. *Nucleic Acids Res* 2019; **47**(11): 5539-49.

15.	Csardi G, Nepusz T. The igraph software. *Complex syst* 2006; **1695**: 1-9.

16.	Matlock W, Lipworth S, Chau KK, et al. Enterobacterales plasmid sharing amongst human bloodstream infections, livestock, wastewater, and waterway niches in Oxfordshire, UK. *eLife* 2023; **12**.

17.	Letunic I, Bork P. Interactive Tree of Life (iTOL) v6: recent updates to the phylogenetic tree display and annotation tool. *Nucleic Acids Res* 2024; **52**(W1): W78-W82.
